# SARS-CoV-2 subgenomic RNA kinetics in longitudinal clinical samples

**DOI:** 10.1101/2021.04.26.21256131

**Authors:** Renu Verma, Eugene Kim, Giovanny Joel Martinez-Colón, Prasanna Jagannathan, Arjun Rustagi, Julie Parsonnet, Hector Bonilla, Chaitan Khosla, Marisa Holubar, Aruna Subramanian, Upinder Singh, Yvonne Maldonado, Catherine A. Blish, Jason R. Andrews

## Abstract

**Background:** Given the persistence of viral RNA in clinically recovered COVID-19 patients, subgenomic RNAs (sgRNA) have been reported as potential molecular viability markers for SARS-CoV-2. However, few data are available on their longitudinal kinetics, compared with genomic RNA (gRNA), in clinical samples.

**Methods:** We analyzed 536 samples from 205 patients with COVID-19 from placebo-controlled, outpatient trials of Peginterferon Lambda-1a (Lambda; n=177) and favipiravir (n=359). Nasal swabs were collected at three time points in the Lambda (Day 1, 4 and 6) and favipiravir (Day 1, 5, and 10) trials. N-gene gRNA and sgRNA were quantified by RT-qPCR. To investigate the decay kinetics *in vitro*, we measured gRNA and sgRNA in A549^ACE2+^ cells infected with SARS-CoV-2, following treatment with remdesivir or DMSO control.

**Results:** At six days in the Lambda trial and ten days in the favipiravir trial, sgRNA remained detectable in 51.6% (32/62) and 49.5% (51/106) of the samples, respectively. Cycle threshold (Ct) values for gRNA and sgRNA were highly linearly correlated (Pearson’s r=0.87) and the rate of increase did not differ significantly in Lambda (1.36 cycles/day vs 1.36 cycles/day; p = 0.97) or favipiravir (1.03 cycles/day vs 0.94 cycles/day; p=0.26) trials. From samples collected 15-21 days after symptom onset, sgRNA was detectable in 48.1% (40/83) of participants. In SARS-CoV-2 infected A549^ACE2+^ cells treated with remdesivir, the rate of Ct increase did not differ between gRNA and sgRNA.

**Conclusions:** In clinical samples and *in vitro*, sgRNA was highly correlated with gRNA and did not demonstrate different decay patterns to support its application as a viability marker.

**Summary:** We observed prolonged detection of subgenomic RNA in nasal swabs and equivalent decay rates to genomic RNA in both longitudinal nasal swabs and in remdesivir-treated A549^ACE2+^ cells infected with SARS-CoV-2. Taken together, these findings suggest that subgenomic RNA from SARS-CoV-2 is comparably stable to genomic RNA and that its detection is therefore not a more reliable indicator of replicating virus.

## INTRODUCTION

Understanding and quantifying the replicating or transcriptionally active virus among individuals with SARS-CoV-2 could inform treatment decisions and response monitoring, as well as the need for isolation, contact tracing and infection control measures. The duration of infectiousness as estimated from transmission studies appears much shorter than the duration of PCR positivity in airway secretions (1,2). Studies comparing culture and RT-PCR from the same samples have revealed that there is often substantial discrepancy between these measurements, with PCR remaining positive for days to weeks longer than culture (3-7). While culture remains the reference standard for detection of infectious virus, it may lack sensitivity, and it requires biosafety level 3 facilities, precluding its use at scale as a clinical or public health tool (8). To overcome this obstacle, there has been considerable interest in the development of molecular viability markers to sensitively detect and quantify transcriptionally active virus (3,9,10).

SARS-CoV-2 is an enveloped, positive sense, single-stranded RNA virus which employs a complicated pattern of replication as well as transcription of genome length and smaller sgRNAs (11). These sgRNAs are transcriptional intermediates, susceptible to enzymatic degradation, and are not believed to be packaged in the final progeny virion, making them an attractive marker for an actively transcribing virus (12). Small clinical studies have suggested that, compared with gRNA, sgRNA correlates better with culturable virus. These studies targeted N-gene to detect gRNA and compared sgRNA stability with a relatively less abundant/sensitive E-gene sgRNA assay (2,3,9). Despite the E-gene sgRNA assay possibly being suboptimal and may give false negative results, these findings have led to the use of sgRNA assay as an outcome in preclinical investigation of novel therapies (13,14) and its suggested use to terminate medical isolation for individuals with COVID-19 (3). In contrast, a recent study found that sgRNAs were detectable up to 17 days after initial detection and that they may be protected from nuclease degradation by double membrane vesicles (15). However, because this study only had 12 clinical samples, further evidence about the kinetics of sgRNA versus gRNA in longitudinal samples is needed to determine whether sgRNA abundance better reflects recently transcribing viral infection. Additionally, in order to serve as a marker of replicating virus, sgRNA is expected to show a rapid decline after transcriptional inhibition due to ribonuclease degradation, in contrast to gRNA, which may be protected from degradation by viral capsids and therefore persist more durably (16). Therefore, we hypothesized that, upon treatment with SARS-CoV-2 RNA-dependent RNA polymerase inhibitors (17, 18, 19) in cell lines infected with SARS-CoV-2, we should observe a rapid decline of sgRNA after viral death when compared with gRNA.

To address these gaps, we developed an N-gene sgRNA assay to directly compare its stability with N-gene gRNA. sgRNAs in SARS-CoV-2 share a common leader sequence at the 5’ end which is absent in the gene amplified from the gRNA (9). We combined the common leader sequence as forward primer with CDC N1 gene assay’s reverse primer (20) to facilitate comparison of N-gene sgRNA with gRNA copies. We applied this assay to serial samples from individuals participating in two randomized clinical trials to characterize decay rates. Additionally, we leveraged the inhibition of viral transcription and replication by an RNA-dependent RNA polymerase inhibitor (remdesivir) (19) to measure and compare the decay kinetics of gRNA and sgRNA following polymerase inhibition in SARS-CoV-2 infected A549^ACE2+^ cells.

## METHODS

### Ethics statement

All participants were >18 years of age and provided written informed consent. The studies were approved by the Stanford IRB (#57686 and #58869).

### Overview and Study Population

This was a sub-study of two Phase 2 randomized, placebo-controlled trials of peginterferon-Lambda-1a (Lambda) (NCT04331899) and favipiravir (NCT04346628) for treatment of COVID-19. Individuals >18 years of age with RT-PCR confirmed SARS-CoV-2 infection were recruited to participate and were eligible if they could be randomized within 72 hours of a positive SARS-CoV-2 test and were not hospitalized. Additional exclusion criteria were respiratory rate < 20 breaths per minute, room air oxygen saturation <94%, pregnancy or breastfeeding, or use of other investigational agents for treatment of COVID-19. In the Lambda trial, enrolled participants were randomized to a single injection with 180 mcg of Lambda versus placebo injection and followed for 28 days. In the favipiravir trial, individuals were randomized to oral favipiravir tablets (1800 mg on day 1, followed by 800 mg twice daily for 9 days) or matching placebo. The primary outcome for both studies was time to cessation of viral shedding as measured by qRT-PCR performed on oropharyngeal swab samples (Lambda trial) or nasal swabs (favipiravir trial). In August 2020, we amended both protocols to collect nasal swabs (LH-11-10 Longhorn Hydra Sterile Flocked Swab) to assay for gRNA and sgRNA. For Lambda trial, it was earlier found that in both patients receiving Lambda and placebo, the median time to cessation of viral shedding was 7 days (21). A single dose of subcutaneous Peginterferon Lambda-1a neither shortened the duration of SARS-CoV-2 viral shedding nor improved symptoms in outpatients (21). The favipiravir trial is an ongoing study and remains blinded.

### RNA extraction and quantitative RT-PCR assay for SARS-CoV-2 RNA

Nasal swabs were collected and transported in 500 ul of Primestore MTM (Longhorn Vaccines & Diagnostics) RNA stabilizing media. RNA was extracted using MagMAX™ Viral/Pathogen Ultra Nucleic Acid Isolation Kit (Cat # A42356 Applied Biosystems) according to the manufacturer’s instructions and eluted in 50ul of elution buffer. We performed qRT-PCR for the N gene using the CDC qualified primers and probes amplifying N1 region of SARS-CoV-2 N-gene (20). TaqPath one-step RT-PCR mastermix (Invitrogen, Darmstadt, Germany) was used in a 20ul reaction volume and the samples were analyzed on a StepOne-Plus (Applied Biosystems) instrument, using the following program: 10 min at 50 °C for reverse transcription, followed by 3 min at 95 °C and 40 cycles of 10 s at 95 °C, 15 s at 56 °C, and 5 s at 72 °C. We estimated copies/sample from a standard curve using a pET21b+ plasmid (GenScript, USA) with the N-gene. The cycle threshold (Ct) cutoff for positive samples was <38.

### Quantitative RT-PCR Assay for SARS-CoV-2 sgRNA

Since all sgRNAs are known to carry a common leader sequence, to amplify N-gene sgRNA, we combined a previously described E-gene sgRNA forward primer for SARS-CoV-2 leader sequence along with the CDC N1-gene segment reverse primer and probe to detect N-gene sgRNA (3). We used TaqPath one-step RT-PCR mastermix with 400 nM concentrations of each of the primer and 200 nM of probe to amplify sgRNA. The N-gene PCR reactions conditions were used for sgRNA amplification. We estimated copies/sample from a standard curve using a pET21b+ plasmid with the N-gene sgRNA sequence. The cycle threshold (Ct) cutoff for positive samples was <38.

### sgRNA validation by Sanger sequencing

For the first 15 positive clinical samples, we confirmed amplification product identity by Sanger sequencing. We performed endpoint PCR using the same primers, purified it by gel electrophoresis, and performed Sanger sequencing with these primers. The resulting sequences were aligned using to SARS-CoV-2 genome (GenBank: MT568638.1) to compare sequence similarity of the product with the leader sequence and N-gene. Samples were considered positive for sgRNA if the leader sequence identity with the reference genome was greater than 98%.

### sgRNA kinetics in SARS-CoV-2 infected A549^ACE2+^ cells

#### Cell culture and in vitro SARS-CoV-2 infection

The human lung epithelial carcinoma cell line, A549, overexpressing Angiotensin-converting enzyme 2 (ACE2), A549^ACE2+^, was provided by Ralf Bartenschlager (Heidelberg University) (22). A549^ACE2+^ cells were cultured in Dulbecco’s Modified Eagle Medium (DMEM) (Life technologies; 11885-092) supplemented with 10% Fetal bovine serum (Corning; MT35016CV), 1% Penicillin-Streptomycin (Thermo Fisher Scientific; 15070063), and 623ug/ml of Geneticin (Thermo Fisher Scientific; 10121035). For viral infection, cells were seeded a day before infection by culturing 1×10^5^ cells per well in a 6-well plate (Corning). Cells were at passage 14 at the time of infection. Viral infection was performed with the Washington strain of SARS-CoV-2 (2019-nCOV/USA-WA1/2020), titered by plaque assay on VeroE6 cells, at a multiplicity of infection (MOI) of 1. Briefly, in Biosafety level 3 (BSL3) containment, culture media was removed, and cells were washed with phosphate buffered saline (PBS) (Thermo Fisher Scientific; 10-296-028) multiple times before adding the viral stock. Cells were then incubated at 37°C with 5% CO_2_ for 1 hour while gently rocking. After 1 hour, cells were washed with 1x PBS and incubated in culture media. Supernatant and cells were collected at 1 and 24 hours post-infection (hpi) in TRIzol LS (Thermo Fisher Scientific; 10010023) for RNA extraction. Other wells were either treated with 0.1% final concentration of dimethylsulfoxide (DMSO) (Sigma Life Science: Cat# D2650) or 10µM remdesivir (Gilead, Cat# NDC 61958-2901-2) in 0.1% DMSO and cultured for longer periods (48, 72, and 96hpi). It has been previously demonstrated that at 10µM prodrug concentration, remdesivir potently inhibits SARS-CoV-2 in A549^ACE2+^ cells (23). Cytopathic effect on SARS-CoV-2 in vitro-infected A549^ACE2+^ cells that were treated with either 10µM remdesivir or vehicle, 0.1% DMSO, was monitored before and after infection. Cell line experiments at all time-points and treatment conditions were performed in technical duplicates. Cells and supernatant were collected, and RNA was extracted independently for all technical duplicates without pooling. An image of cells was collected using an EVOS XL core imaging system (Thermo Fisher scientific), with a 10x objective, before collecting cell pellet and supernatant from each treatment, and time point.

#### RNA extraction and RT-PCR

RNA from supernatant and cells collected at 1, 24, 48, 72 and 96 hpi in TRIzol LS was extracted using isolated using standard phenol-chloroform extraction per manufacturer’s instructions. The SARS-CoV-2 genomic and sgRNA RT-qPCR assays from cell line technical duplicates were further performed in technical duplicates. The RNA copies were quantified using standard curves derived from plasmids. Eukaryotic 18S rRNA commercial TaqMan assay (4333760T, Thermo Fisher Scientific) was used as an internal control.

### Statistical Analyses

We estimated the change in cycle threshold (Ct) value for gRNA and sgRNA by day using generalized linear mixed models with random effect for participant. We tested for differences in the coefficients for collection day for outcomes of gRNA and sgRNA by performing ANOVA on a joint model with a dummy variable for RNA type. We used generalized additive mixed models with a random effect for participant to investigate the relationship between sample collection day and cycle threshold (Ct) values. All analyses were performed using R (24).

## RESULTS

### Study population characteristics

We recruited 205 COVID-19 positive patients from placebo-controlled trials of interferon Lambda (n=66) and Favipiravir (n=139) between August, 2020 and January, 2021. All participants were enrolled in the trials within 72 hours of a positive SARS-CoV-2 RT-qPCR test. Median age of the participants was 40 years (range, 18-73), and 46.8% (96/205) were female. The majority of participants (197/205; 96.1%) reported one or more COVID-19 related symptoms several days prior to enrollment (median, 5 days; range, 0-21). Symptoms with onset more than three weeks prior to study enrollment were not considered to be associated with COVID-19. The most common baseline symptoms reported by the patients before randomization were cough, diarrhea, body ache, headache, fatigue and shortness of breath (Table 1).

**Table 1:** Characteristics of the study participants recruited from the Lambda and favipiravir trials.

|  | Trial |  |  |
| --- | --- | --- | --- |
|  | Lambda (n=66) | Favipiravir (n=139) | Overall (n=205) |
| **Age in years, median (IQR)** | 36 (31-48.5) | 42 (33-53) | 40 (32-52) |
| **Female, n (%)** | 28 (47.5%) | 60 (46.2%) | 88 (46.6%) |
| **Symptomatic at enrollment, n (%)** | 66 (100%) | 131 (94.2%) | 197 (96.1%) |
| **Duration of symptoms prior to enrollment, days, median (IQR)** | 5 (4-7.75) | 5 (3-7) | 5 (3-7) |
| 0-7 days, n (%) | 49 (74.2%) | 110 (79.1%) | 159 (77.6%) |
| 8-14 days, n (%) | 14 (21.2%) | 20 (14.4%) | 34 (16.6%) |
| 15-21 days, n (%) | 3 (4.5%) | 1 (0.7%) | 4 (2.0%) |

### gRNA and sgRNA RT-qPCR positivity in clinical samples

We analyzed 536 nasal swab samples collected from 205 COVID-19 patients from the Lambda (n=177) and favipiravir (n=359) trials between 0 to 21 days post symptom onset. For the favipiravir trial, nasal swabs were collected on the day of enrollment (day 1), followed by day 5 and day 10. For Lambda trial, nasal swabs were collected on the day of enrollment (day 1) followed by day 4 and day 6. Overall gRNA RT-qPCR positivity in samples from the favipiravir trial on day 1, 5 and 10 was 91.5%, 82.9% and 60.3% respectively (Table 2). For sgRNA, positivity was 89.2%, 77.2% and 49.5%. For the Lambda trial, overall gRNA positivity on day 1, 4 and 6 was 91.6%, 90.9% and 91.9% respectively. For sgRNA, overall positivity was 81.6%, 74.5% and 51.6%. We observed a high correlation (Pearson’s r= 0.87) between the cycle threshold (Ct) values of gRNA and sgRNA at all time points, and detection of sgRNA was strongly predicted by gRNA Ct (**Figure 1**). For the first 15 samples for which sgRNA showed positive amplification, we performed Sanger sequencing. All fifteen samples had more than 98% identity with SARS-CoV-2 leader sequence, confirming amplification of the sgRNA transcript (Supplementary figure 1). In a subset of 35 samples in which we performed testing for E-gene sgRNA using a previously published assay, we found high correlation (Pearson’s r=0.89) with N-gene sgRNA, but with higher Ct values (median difference, 4.1 cycles) among positive samples. Among 35 samples positive for N-gene sgRNA, 69.0% (24/35) were negative for E-gene sgRNA (Supplementary figure 2).

**Table 2:** Genomic and sgRNA RT-qPCR positivity in longitudinal samples from the Lambda and favipiravir trials.

| Longitudinal swab samples from Lambda clinical trial (n=177) |  |  |  |  |  |  |
| --- | --- | --- | --- | --- | --- | --- |
|  | Day 1 |  | Day 4 |  | Day 6 |  |
|  | Positive, n (%) | Median Ct | Positive (%) | Median Ct | Positive (%) | Median Ct |
| **gRNA** | 55/60 (91.6%) | 24.4 | 50/55 (90.9%) | 30.2 | 57/62 (91.9%) | 33.2 |
| **sgRNA** | 49/60 (81.6%) | 26.9 | 41/55 (74.5%) | 32.3 | 32/62(51.6%) | 37.3 |
| Longitudinal swab samples from favipiravir clinical trial (n=359) |  |  |  |  |  |  |
|  | Day 1 |  | Day 5 |  | Day 10 |  |
|  | Positive (%) | Median Ct | Positive (%) | Median Ct | Positive (%) | Median Ct |
| **gRNA** | 119/130 (91.5%) | 23.8 | 102/123 (82.9%) | 29.4 | 64/106<br>(60.3%) | 34.4 |
| **sgRNA** | 116/130 (89.2%) | 25.4 | 95/123 (77.2%) | 31.0 | 51/106 (49.5<br>%) | 38 |

Ct cutoff value for positive samples <38; Ct: cycle threshold

**Figure 1:**
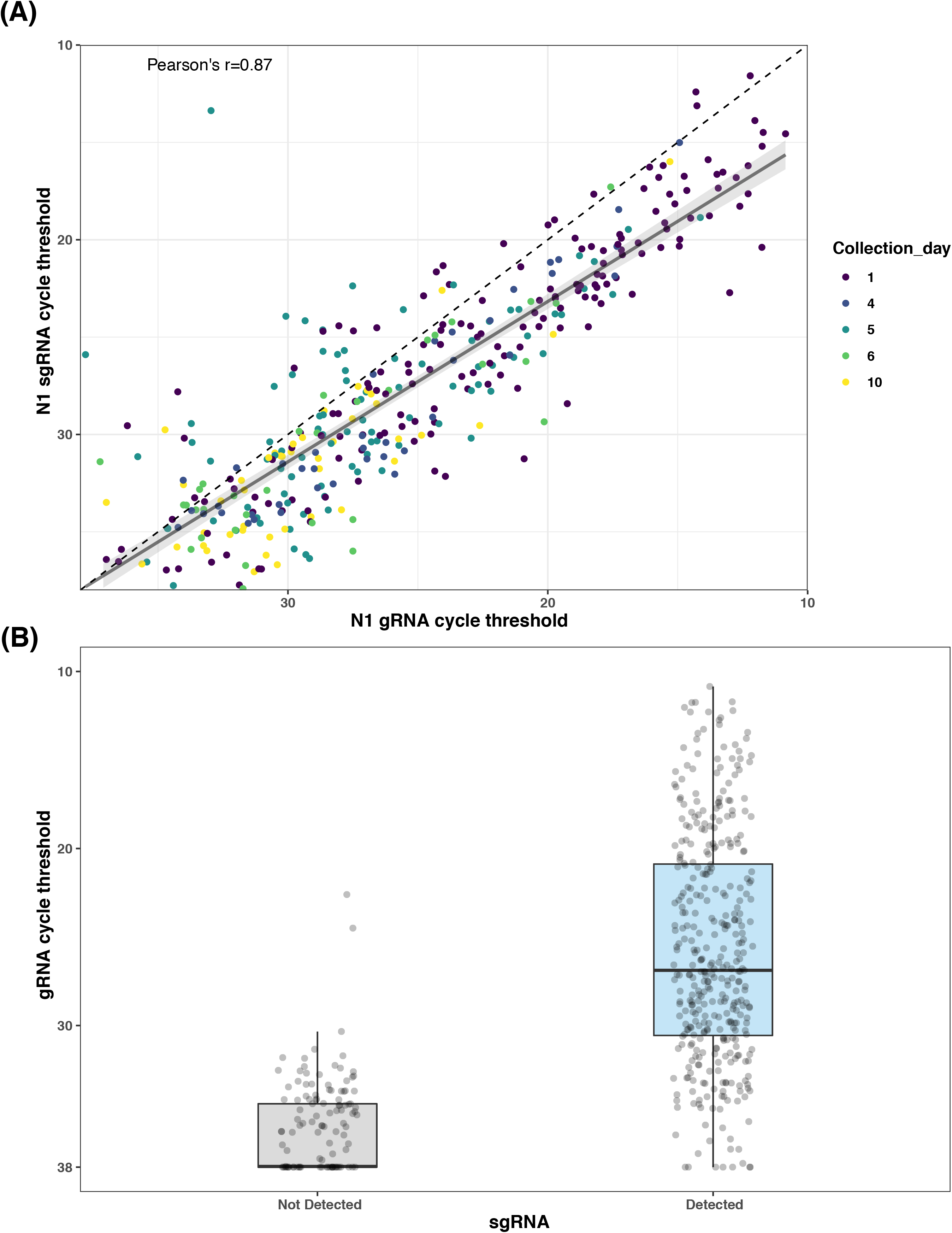
Correlation between genomic and subgenomic RNA in clinical samples. **(A)** Genomic and sgRNA Ct values from clinical samples showed strong correlation (Pearson’s r= 0.87). Dashed line reflects the diagonal of equal cycle thresholds, and solid line is the best fit regression line with shaded area indicating the 95% confidence interval. **(B)** Detection of sgRNA was predicted by cycle threshold value of gRNA.

Randomization data was available for the Lambda study while favipiravir still remains blinded. We did not observe any significant difference in the Ct values of sgRNA between Lambda and placebo recipients. At day six, the gRNA percentage positivity was 83.8% (26/31) (median Ct value = 32.5) in Lambda and 100% (31/31) (median Ct value =33.4) in placebo arm. In sgRNA at day six, percentage positivity was 48.3% (15/31) (median Ct value = 38.0) in Lambda and 54.8% (17/31) (median Ct value =35.9) in placebo arm (p=0.903).

We found no difference in the rate of Ct value increase by day in gRNA compared with sgRNA in the Lambda (1.36 cycles/day vs 1.36 cycles/day; p=0.97) or favipiravir (1.03 cycles/day vs 0.94 cycles/day; p=0.26) trials (Figure 2). Among samples collected 15-21 days after symptom onset from both trials combined, sgRNA was detectable in 48.1% (40/83) of participants (Figure 3).

**Figure 2:**
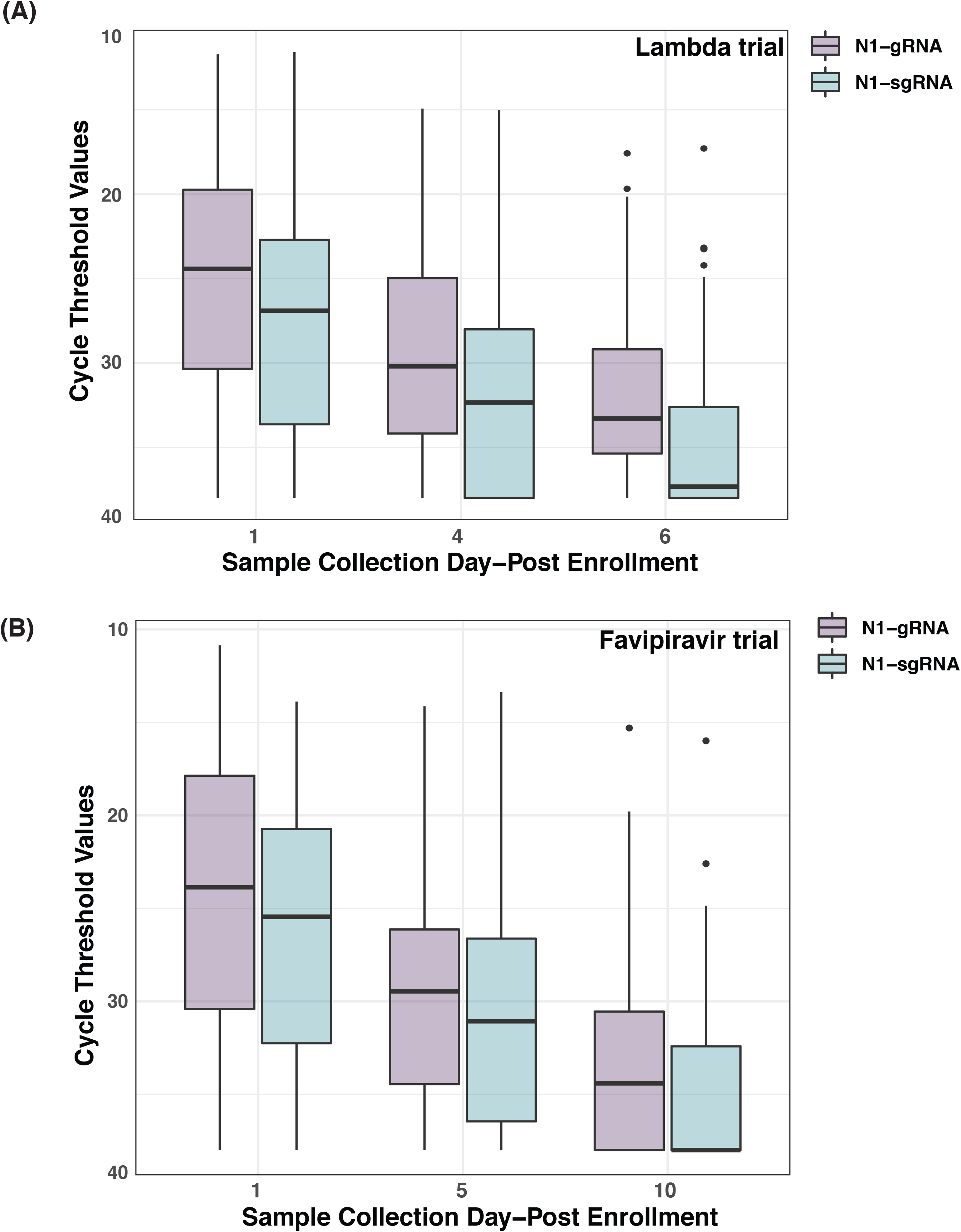
Cycle threshold values for genomic and subgenomic RNA by sample collection day. Boxplot representing Ct values for genomic (Purple) and sgRNA (Blue) from serially collected COVID-19 samples. **(A**) N-gene genomic and sgRNA decay trend in samples from the Lambda trial collected at day 1, 4 and 6. **(B)** N-gene genomic and sgRNA decay trend in samples from the favipiravir trial collected at day 1, 5 and 10.

**Figure 3:**
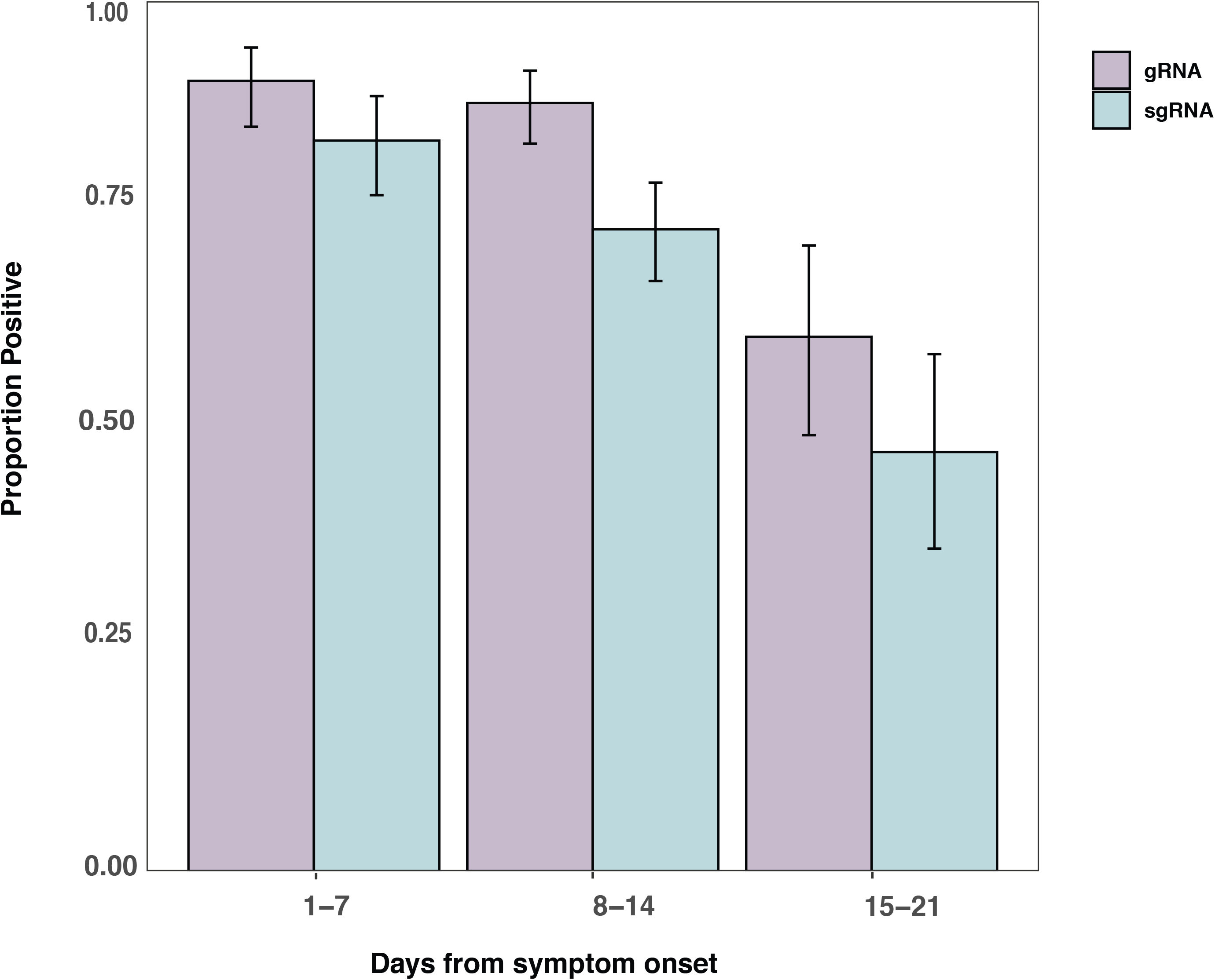
Proportion of samples positive for genomic and subgenomic RNA from time of symptom onset. Samples from the Lambda and favipiravir trials were combined, and a positive sample was one with cycle threshold <38. Error bars denote 95% exact binomial confidence intervals.

### sgRNA kinetics in SARS-CoV-2 infected A549^ACE2+^cells treated with remdesivir

We compared SARS-CoV-2 gRNA and sgRNA degradation kinetics after transcriptional inhibition by antiviral drug remdesivir. We treated SARS-CoV-2 infected A549^ACE2+^cells with 0.1% DMSO vehicle control and 10µM remdesivir at 24hpi. Cytopathic effects were observed in cells treated with DMSO control but not remdesivir (Supplementary figure 3). Compared with DMSO treated cells, SARS-CoV-2 replication in remdesivir treated cells was markedly reduced (nadir gRNA Ct 9.6 vs 14.2; nadir sgRNA Ct 10.0 vs 14.5). In remdesivir-treated cells, gRNA and sgRNA Ct values rose at similar rates in cells (0.11/hour vs 0.09/hour; p=0.153) and declined by similar rates in supernatant (−0.06/hour vs 0.06/hour; p=0.914) (Figure 4).

**Figure 4:**
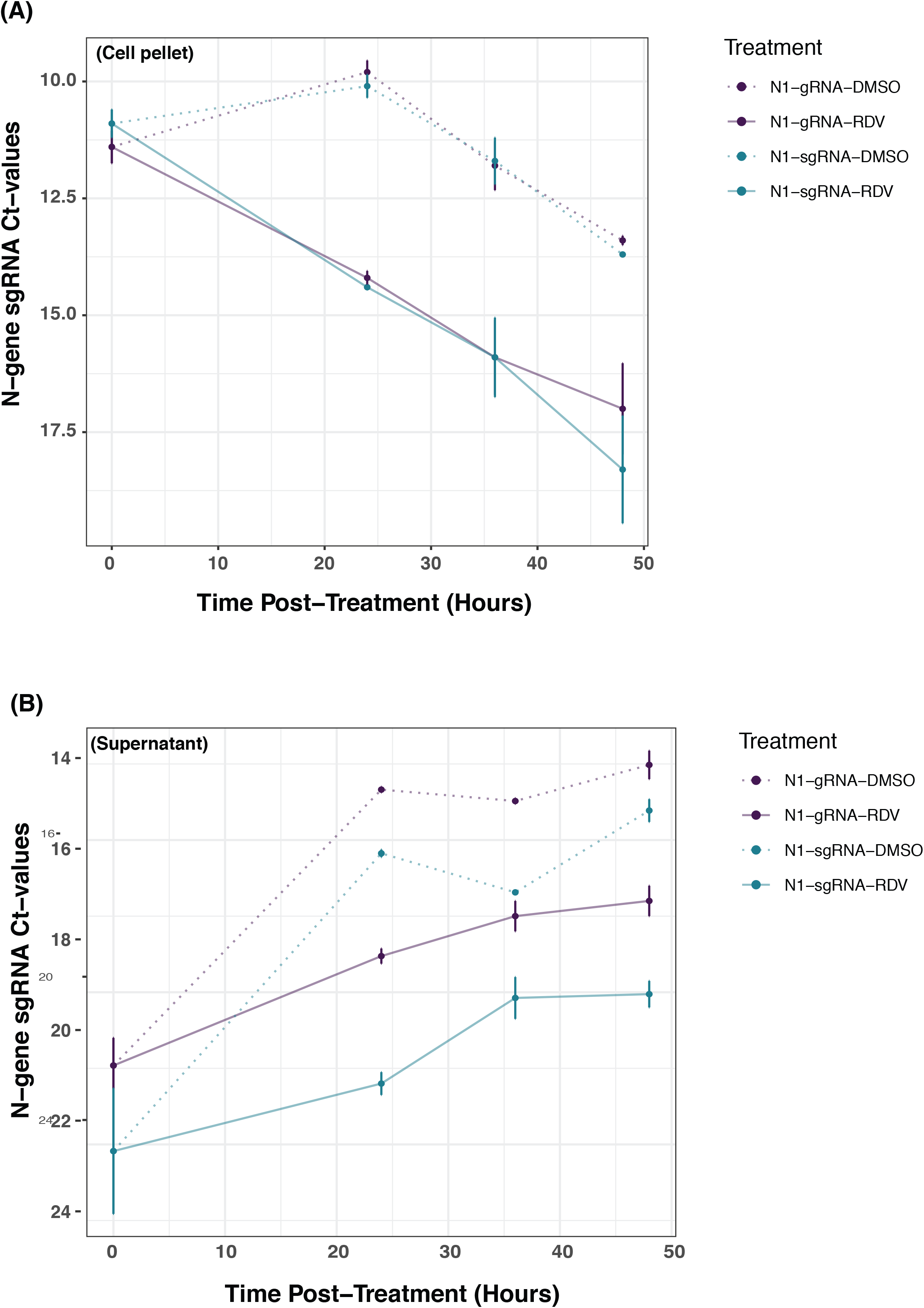
SARS-CoV-2 genomic and subgenomic RNA kinetics in cell culture. SARS-CoV-2 genomic and sgRNA kinetics in A549^ACE2+^ cells treated with 0.1% DMSO and 10µM remdesivir (RDV). Solid lines indicate genomic (Purple) and sgRNA (Blue) levels in A549^ACE2+^ cells treated with remdesivir. Dotted lines indicate genomic (Purple) and sgRNA (Blue) levels in cells treated with 0.1% DMSO. (A) Corresponds to genomic and sgRNA degradation kinetics in washed cell pellets from A549^ACE2+^ cell line. (B) Corresponds to genomic and sgRNA degradation kinetics in supernatant from A549^ACE2+^ cell line.

## DISCUSSION

While there has been considerable interest in the use of sgRNAs as markers of replicating SARS-CoV-2 infection, evidence concerning the decay of sgRNA following onset of infection in humans and cell culture has been lacking. Using longitudinal samples from two clinical trials, we found that sgRNA was detectable in 46% of participants from the Lambda trial and 50% from favipiravir trial 15-21 days after symptoms onset. While gRNA was detectable for longer than sgRNA, they were highly correlated and had indistinguishable rates of decline within individuals over time. We found consistent results in cell culture, whereby gRNA and sgRNA copies declined at the same rate following inhibition of transcription by remdesivir. Taken together, these findings suggest that detection of sgRNAs is not a reliable marker of recent viral transcription and does not provide marginal information over quantification of gRNA. Earlier findings of greater specificity of sgRNAs than gRNAs compared with a reference standard of culture may be explained by the lower analytical sensitivity of the sgRNA assays (3,9,10), particularly using less sensitive E-gene assays.

RNA transcripts have been used as markers of viability or metabolic activity for a number of bacterial and viral pathogens. In bacteria, mRNAs have much shorter half-lives than DNA due to degradation by ribonucleases, such that their presence indicates recent metabolic activity (25,26,27). Similarly, RNA transcription assays have been used to assess replication competent viral pool size for HIV-1 (28). For SARS-CoV-2, sgRNA transcription is believed to occur inside double membrane vesicles, which may protect viral genomic and subgenomic RNA from cytoplasmic degradation due to host enzymes (29-33). We found that sgRNA and gRNA increased at the same rate following infection of cells and then declined at the same rate following cell death (in the control cells treated with DMSO) or following inhibition of RDRP (RNA dependent RNA polymerase) by remdesivir. If sgRNAs were rapidly degraded by ribonucleases, we would have expected a more rapid decline in sgRNAs compared with gRNAs, but this was not observed. Similarly, in the supernatant, we saw no difference in change in sgRNAs compared with gRNAs, again failing to identify rapid clearance of sgRNAs by ribonucleases.

Several previous studies have targeted E-gene sgRNA, reporting in small clinical series that these correlated well with culture (34, 35). This finding may be explained by the fact that E-gene transcripts are less abundant than N-gene transcripts, so assays targeting them will have lower analytical sensitivity (36). Indeed, we performed direct comparison of N-gene and E-gene assays and found that the latter were approximately 5 Ct values higher for the same sample. There is not clear premise to infer that E-gene transcripts are more rapidly degraded by ribonucleases or that they better reflect recent transcription than N-gene sgRNAs. Negative-strand RNA detection in SARS-CoV-2 has recently been reported as another potential viability marker (37). In a study by Alexanderson and group, negative strand RNA was detected up to 11 days (15). However, negative strand assays may be less analytically sensitive than sgRNA assays (37). This could make them appear to be more specific, compared with culture, in clinical samples as they are more likely to be negative when viral abundance is low.

Our study has several limitations, which include lack of viral culture data and samples at later timepoints. Additionally, to study the sgRNA decay kinetics, we targeted a single and highly abundant gene to compare and quantify gRNA and sgRNA, though we found high correlation between E-gene and N-gene sgRNA copy numbers. Analyzing additional targets would help us understand the stability of other sgRNAs. Another limitation our study is lack of unblinded data from the favipiravir study, precluding analysis by study arm. However, our findings of sgRNA persistence in clinical samples and lack of difference in its decline compared with gRNA are notable regardless of whether favipiravir reduced levels in one arm. For remdesivir, a RDRP inhibitor like favipiravir, we found no differential effect on sgRNA, compared with gRNA, in cell culture, despite effective reduction in viral replication and cytopathic effects.

In summary, we found that SARS-CoV-2 sgRNAs are persistently detectable in clinical samples, correlate strongly with gRNA, and decline at indistinguishable rates in clinical samples and cell culture. We find little evidence to support the premise that sgRNA detection is a reliable marker of transcriptionally active virus or that it provides additional information beyond detection of gRNA in clinical samples. We advise caution against using sgRNA assays to inform decisions concerning treatment or medical isolation.

## Data Availability

Data generated in the current study and used for the figures is provided in the manuscript or supplementary files. Additional information can be provided by the Corresponding Author upon request.

## Acknowledgements

We thank the study teams and participants of the Lambda and favipiravir clinical trials. SARS-Related Coronavirus 2, Isolate USA-WA1/2020, NR-52281 was deposited by the CDC and obtained through BEI Resources, NIAID, NIH. We thank Jaishree Garhyan, Director, Invitro BSL-3 Service Center, Stanford School of Medicine.

## Author contributions

JRA and RV conceived the idea of the study. JRA, RV, GMC and CB designed the experiments. PJ, HB, US, AS, MH, YM, and JRA enrolled the clinical cohorts. RV, EK, and GMC performed the experiments. JRA, RV and EK analyzed data. RV and JRA wrote the first draft of the manuscript, and all authors contributed to the final version.

## Figures

**Supplementary Figure 1:**
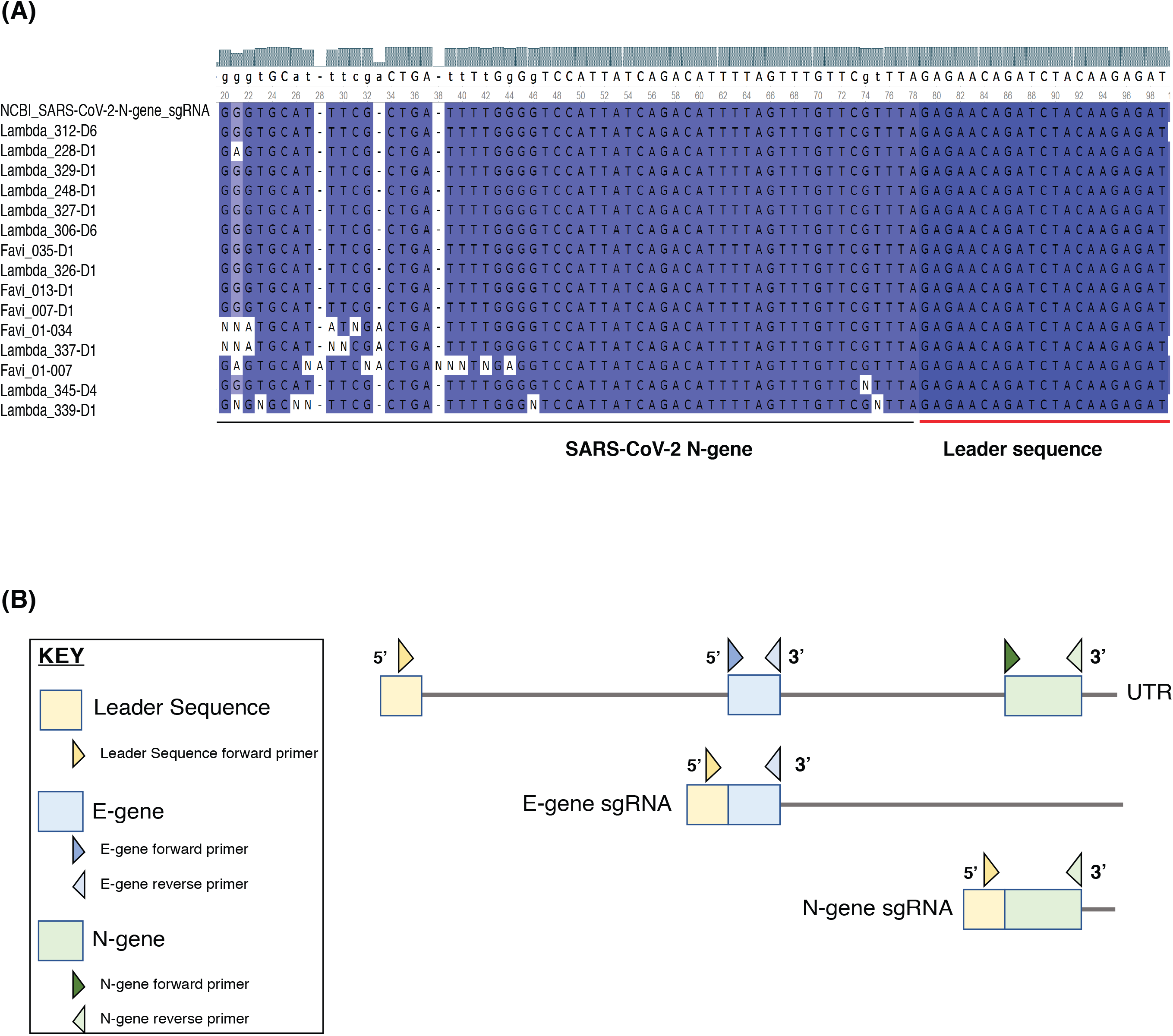
(A) Sanger sequencing validation: SARS-CoV-2 sgRNA RT-PCR results confirmed on Sanger sequencing. The samples were mapped to SARS-CoV-2 genome (GenBank: MT568638.1) using MUSCLE (MUltiple Sequence Comparison by Log-Expectation). Clinical samples assayed for sgRNA mapped to the leader sequence and N-gene. **(B)** Schema on sgRNA synthesis in SARS-CoV-2. In full-length genomic RNA, the 5’ terminal contains a leader sequence (yellow). During viral transcription, leader sequence independently fuses with different genes, resulting in smaller sgRNAs containing a common leader sequence.

**Supplementary Figure 2:**
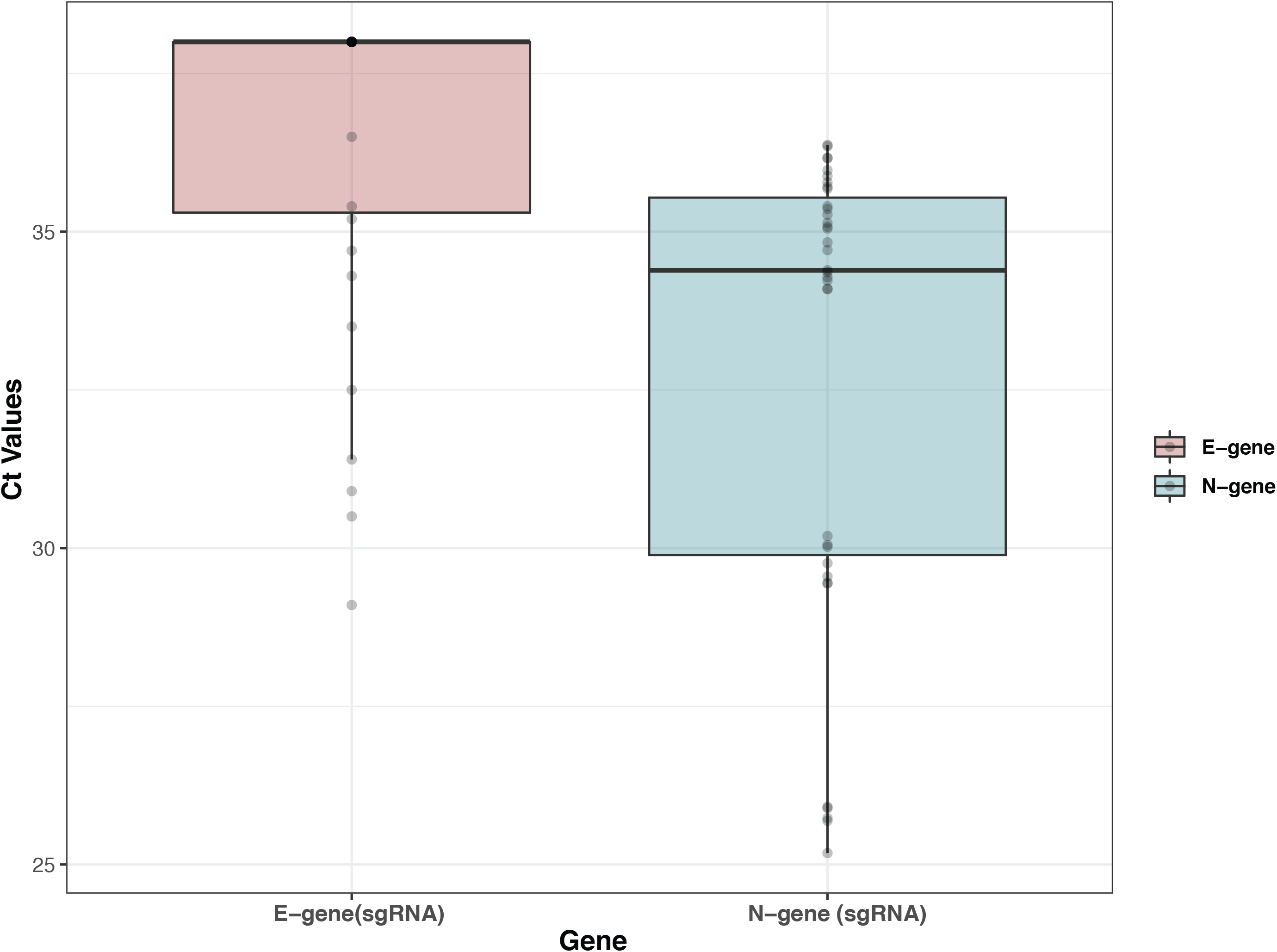
Boxplot representing Ct values for N-gene sgRNA (Blue) and E-gene sgRNA (Pink). Higher Ct values were observed for the E-gene sgRNA among the samples tested positive for N-gene sgRNA. A large fraction of samples 69% (24/35), which were positive for N-gene sgRNA were negative for E-gene sgRNA suggesting poor assay sensitivity.

**Supplementary Figure 3:**
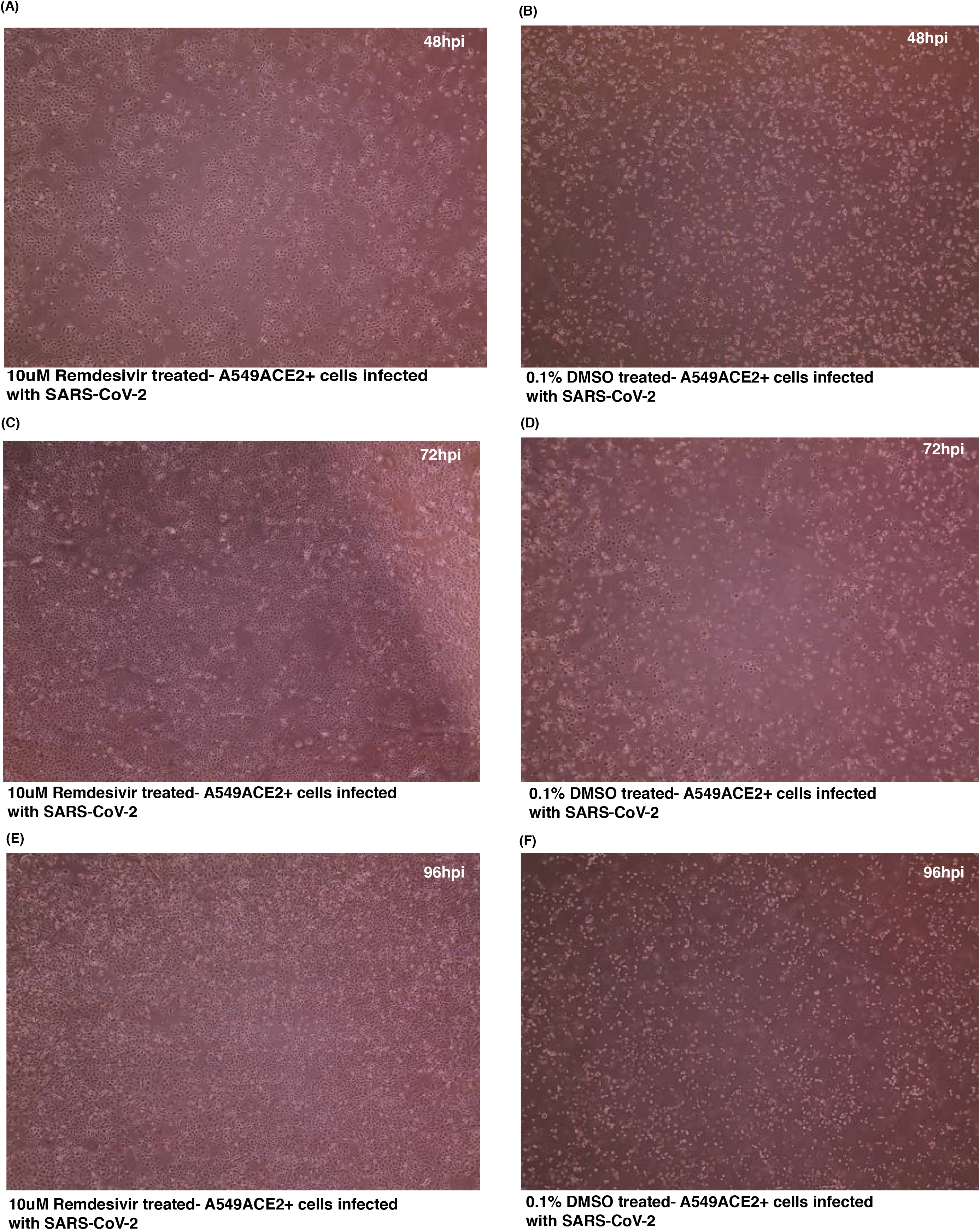
Cytopathic effect of SARS-CoV-2 on A549^ACE2+^ cells treated with either 10µM remdesivir or 0.1% DMSO. The images were collected using an EVOS XL core imaging system (Thermo Fisher scientific), with a 10x objective, before collecting cell pellet and supernatant from each treatment and time point. Cytopathic effect was characterized by significant changes in morphology from well-spread epithelial cells to small rounded cells, the inability to achieve confluency, and presence of excessive and unusual rounded floating cells, a characteristic of cell death. (A) Remdesivir treated A549^ACE2+^cells 24hours after treatment at 48hpi (B) DMSO treated A549^ACE2+^cells 24hours after treatment at 48hpi (C) Remdesivir treated A549^ACE2+^cells 48hours after treatment at 72hpi (D) DMSO treated A549^ACE2+^cells 48hours after treatment at 72hpi (E) Remdesivir treated A549^ACE2+^cells 72hours after treatment at 96hpi (F) DMSO treated A549^ACE2+^cells 72hours after treatment at 96hpi.

**Supplementary Table 1**: RT-qPCR data on N-gene genomic and sgRNA degradation analysis in remdesivir and DMSO-treated A549^ACE2+^ cells. The cells were infected with SARS-CoV-2 at 24, 48hpi, 72hpi and 96hpi. Total number of copies per samples corresponding to Ct values were calculated from standard curves using a standard curve derived from pET21b+ plasmids with the target insert.

