## supplementary table_1 for "SARS-CoV-2 subgenomic RNA kinetics in longitudinal clinical samples"

**Supplementary table 1: qPCR results for Remdesivir and DMSO treated cells infected with SARS-CoV-2**

| Sample ID | Sample type | N-gene Ct | N-gene copies | sgRNA N1 Ct | sgRNA N1 copies | 18s rRNA Ct |
|---|---|---|---|---|---|---|
| 1hpi_duplicate_1 | cell | 24.8 | 5.20E+04 | 26.6 | 2.21E+03 | 11.6 |
| 1hpi_duplicate_1 | cell | 24.7 | 5.66E+04 | 26.6 | 2.21E+03 | 11.3 |
| 1hpi_duplicate_2 | cell | 26 | 2.68E+04 | 27.9 | 8.54E+02 | 13.2 |
| 1hpi_duplicate_2 | cell | 25.9 | 2.72E+04 | 28.1 | 7.18E+02 | 13.2 |
| 24hpi_duplicate_1 | cell | 11.4 | 1.05E+08 | 10.9 | 2.85E+08 | 7.1 |
| 24hpi_duplicate_1 | cell | 11.5 | 1.02E+08 | 11 | 2.66E+08 | 6.9 |
| 24hpi_duplicate_2 | cell | 12.1 | 7.11E+07 | 11.4 | 1.93E+08 | 7 |
| 24hpi_duplicate_2 | cell | 12.2 | 6.78E+07 | 11.6 | 1.68E+08 | 7.3 |
| 48hpi-DMSO_duplicate_1 | cell | 9.8 | 2.68E+08 | 10.1 | 5.10E+08 | 8 |
| 48hpi-DMSO_duplicate_1 | cell | 9.9 | 2.55E+08 | 10.4 | 4.12E+08 | 7.8 |
| 48hpi-DMSO_duplicate_2 | cell | 9.3 | 3.49E+08 | 9.7 | 7.11E+08 | 7.1 |
| 48hpi-DMSO_duplicate_2 | cell | 9.4 | 3.42E+08 | 9.7 | 6.91E+08 | 7.1 |
| 48hpi-Remdesivir_duplicate_1 | cell | 14.2 | 2.20E+07 | 14.4 | 2.11E+07 | 8.1 |
| 48hpi-Remdesivir_duplicate_1 | cell | 14.1 | 2.27E+07 | 14.5 | 1.95E+07 | 8.1 |
| 48hpi-Remdesivir_duplicate_2 | cell | 14.5 | 1.87E+07 | 14.5 | 1.86E+07 | 6.9 |
| 48hpi-Remdesivir_duplicate_2 | cell | 14.4 | 1.94E+07 | 14.6 | 1.75E+07 | 7.1 |
| 72hpi-DMSO_duplicate_1 | cell | 11.8 | 8.71E+07 | 11.7 | 1.56E+08 | 8.5 |
| 72hpi-DMSO_duplicate_1 | cell | 11.6 | 9.30E+07 | 11.7 | 1.57E+08 | 8.4 |
| 72hpi-DMSO_duplicate_2 | cell | 10.7 | 1.63E+08 | 10.7 | 3.24E+08 | 7.7 |
| 72hpi-DMSO_duplicate_2 | cell | 10.6 | 1.70E+08 | 10.8 | 2.99E+08 | 7.9 |
| 72hpi-Remdesivir_duplicate_1 | cell | 15.9 | 8.04E+06 | 15.9 | 6.56E+06 | 6.4 |
| 72hpi-Remdesivir_duplicate_1 | cell | 15.8 | 8.52E+06 | 16 | 6.40E+06 | 6.5 |
| 72hpi-Remdesivir_duplicate_2 | cell | 17.5 | 3.24E+06 | 17.7 | 1.79E+06 | 8.3 |
| 72hpi-Remdesivir_duplicate_2 | cell | 17.4 | 3.47E+06 | 17.5 | 2.03E+06 | 8.3 |
| 96hpi-DMSO_duplicate_1 | cell | 13.4 | 3.36E+07 | 13.7 | 3.48E+07 | 9 |
| 96hpi-DMSO_duplicate_1 | cell | 13.5 | 3.19E+07 | 13.7 | 3.46E+07 | 9 |
| 96hpi-DMSO_duplicate_2 | cell | 13.7 | 2.95E+07 | 13.8 | 3.32E+07 | 9.2 |
| 96hpi-DMSO_duplicate_2 | cell | 13.6 | 3.02E+07 | 13.7 | 3.40E+07 | 9.2 |
| 96hpi-Remdesivir_duplicate_1 | cell | 17 | 4.47E+06 | 18.3 | 1.09E+06 | 6.6 |
| 96hpi-Remdesivir_duplicate_1 | cell | 17 | 4.46E+06 | 18.1 | 1.32E+06 | 6.5 |
| 96hpi-Remdesivir_duplicate_2 | cell | 19 | 1.38E+06 | 20.5 | 2.13E+05 | 7.8 |
| 96hpi-Remdesivir_duplicate_2 | cell | 18.9 | 1.46E+06 | 20.5 | 2.23E+05 | 7.9 |
| 1hpi_duplicate_1 | Supernatant | 25.4 | 3.63E+04 | 27.5 | 1.10E+03 | 20.7 |
| 1hpi_duplicate_1 | Supernatant | 25.4 | 3.71E+04 | 27.5 | 1.14E+03 | 20.7 |
| 1hpi_duplicate_2 | Supernatant | 23.1 | 1.39E+05 | 24 | 1.59E+04 | 17.6 |
| 1hpi_duplicate_2 | Supernatant | 23.4 | 1.18E+05 | 23.9 | 1.67E+04 | 17.7 |
| 24hpi_duplicate_1 | Supernatant | 22.7 | 1.77E+05 | 25.9 | 3.88E+03 | 21.1 |
| 24hpi_duplicate_1 | Supernatant | 22.6 | 1.79E+05 | 25.8 | 3.99E+03 | 21.1 |
| 24hpi_duplicate_2 | Supernatant | 21.2 | 3.97E+05 | 22.5 | 4.77E+04 | 19.8 |
| 24hpi_duplicate_2 | Supernatant | 21.2 | 3.97E+05 | 22.5 | 4.74E+04 | 19.9 |
| 48hpi-DMSO_duplicate_1 | Supernatant | 14.8 | 1.58E+07 | 16.5 | 4.39E+06 | 13.4 |
| 48hpi-DMSO_duplicate_1 | Supernatant | 14.7 | 1.63E+07 | 16.4 | 4.53E+06 | 13.4 |

| | | | | | | |
|---|---|---|---|---|---|---|
| 48hpi-DMSO_duplicate_2 | Supernatant | 14.6 | 1.76E+07 | 16.3 | 4.90E+06 | 13.3 |
| 48hpi-DMSO_duplicate_2 | Supernatant | 14.6 | 1.70E+07 | 16.2 | 5.25E+06 | 13.4 |
| 48hpi-Remdesivir_duplicate_1 | Supernatant | 18.8 | 1.59E+06 | 22.1 | 6.61E+04 | 16.8 |
| 48hpi-Remdesivir_duplicate_1 | Supernatant | 18.9 | 1.51E+06 | 22.1 | 6.33E+04 | 16.7 |
| 48hpi-Remdesivir_duplicate_2 | Supernatant | 19.3 | 1.22E+06 | 22.7 | 4.05E+04 | 17.2 |
| 48hpi-Remdesivir_duplicate_2 | Supernatant | 19.2 | 1.25E+06 | 22.7 | 4.09E+04 | 17.3 |
| 72hpi-DMSO_duplicate_1 | Supernatant | 14.9 | 1.48E+07 | 17.4 | 2.27E+06 | 14.6 |
| 72hpi-DMSO_duplicate_1 | Supernatant | 14.9 | 1.45E+07 | 17.4 | 2.26E+06 | 14.6 |
| 72hpi-DMSO_duplicate_2 | Supernatant | 15.1 | 1.34E+07 | 17.3 | 2.37E+06 | 14.5 |
| 72hpi-DMSO_duplicate_2 | Supernatant | 15 | 1.38E+07 | 17.4 | 2.25E+06 | 14.5 |
| 72hpi-Remdesivir_duplicate_1 | Supernatant | 17.5 | 3.33E+06 | 19.6 | 4.10E+05 | 15 |
| 72hpi-Remdesivir_duplicate_1 | Supernatant | 17.7 | 3.02E+06 | 19.6 | 4.17E+05 | 14.9 |
| 72hpi-Remdesivir_duplicate_2 | Supernatant | 18.3 | 2.06E+06 | 20.7 | 1.81E+05 | 15.7 |
| 72hpi-Remdesivir_duplicate_2 | Supernatant | 18.5 | 1.86E+06 | 20.7 | 1.84E+05 | 15.7 |
| 96hpi-DMSO_duplicate_1 | Supernatant | 13.6 | 2.99E+07 | 15 | 1.36E+07 | 13.4 |
| 96hpi-DMSO_duplicate_1 | Supernatant | 13.7 | 2.91E+07 | 14.8 | 1.56E+07 | 13.4 |
| 96hpi-DMSO_duplicate_2 | Supernatant | 14.4 | 1.91E+07 | 15.5 | 9.05E+06 | 14 |
| 96hpi-DMSO_duplicate_2 | Supernatant | 14.4 | 1.97E+07 | 15.6 | 8.75E+06 | 14 |
| 96hpi-Remdesivir_duplicate_1 | Supernatant | 17.1 | 4.07E+06 | 19.7 | 3.88E+05 | 13.3 |
| 96hpi-Remdesivir_duplicate_1 | Supernatant | 17.3 | 3.77E+06 | 19.7 | 3.89E+05 | 13.3 |
| 96hpi-Remdesivir_duplicate_2 | Supernatant | 18 | 2.47E+06 | 20.4 | 2.27E+05 | 13.9 |
| 96hpi-Remdesivir_duplicate_2 | Supernatant | 18 | 2.47E+06 | 20.4 | 2.30E+05 | 14.1 |
